# Effects of Exposure to Forest Fire Smoke on Respiratory Health in Rural Communities of San José de Chiquitos, Bolivia

**DOI:** 10.1101/2025.07.06.25327500

**Authors:** E Toledo German, BA Rodriguez-Olguin, BN Mercado-Saavedra

## Abstract

Forest fires in Bolivia have increased in frequency and intensity, posing environmental and human health threats. This study evaluated the impact of air pollution from forest fires on the health of San José de Chiquitos, Santa Cruz, Bolivia’s population. A cross-sectional study was conducted in two affected communities (Los Ciros and Pororó), collecting epidemiological and environmental data in September 2024. The results show that the symptoms most reported by the population exposed to smoke were respiratory (difficulty breathing in 71.4% of cases), followed by irritation in mucous membranes (77.1%), throat (80.0%), and eyes (85.7%). Although no significant differences were observed in all conditions, Los Ciros had a higher prevalence of symptoms, suggesting that poorer baseline health status could aggravate the effects of smoke. The most frequent mitigation measures were the use of masks (63.6%), staying indoors (54.5%), and the use of wet cloths (21.2%), without finding a significant association between these strategies and the reduction of symptoms. This study highlights the need to improve air pollution protection strategies in communities affected by wildfires and develop prevention policies to mitigate their effects on public health.

## INTRODUCTION

Forest burning is a common practice in different regions of Bolivia, especially in the department of Santa Cruz, where its frequency has increased due to natural and anthropic factors ^1,2^. This situation produces atmospheric pollutants, such as PM2.5 and PM10 particles, carbon monoxide, and other toxic gases, which directly impact air quality and the health of nearby residents ^3^. The consequences include an increase in respiratory and cardiovascular diseases, as well as an increase in premature mortality in exposed populations ^4^. Araujo T. et al. analyzed how forest fires alter soils’ physical and chemical properties in the Chiquitania region, Santa Cruz, Bolivia, in 2019 ^5^. The authors highlight that fires cause significant changes in soil structure, organic matter, and nutrient availability. These findings are relevant to our research, as soil degradation can indirectly influence human health. For example, loss of vegetation cover and soil erosion can increase the dispersion of airborne particles, exacerbating air pollution during and after fires. In addition, declining soil quality can affect food security and water quality, which are essential for the health of local communities.

In October 2024, the situation of forest fires in the department of Santa Cruz was alarming, with more than 50 active fires distributed in several municipalities, including San José de Chiquitos, one of the most affected areas ^6–8^. These fires have devastated more than 7 million hectares of forest, generating serious consequences for the community members’ flora, fauna, and human health. San José de Chiquitos, along with Concepción, Urubichá, and Ascensión de Guarayos, is among the municipalities with the most critical outbreaks, which has mobilized hundreds of firefighters to try to control the fire ^6,7,9^. In addition to environmental losses, the fires have led to health problems in nearby communities, where respiratory and ophthalmic conditions are common due to smoke inhalation and air pollution. Despite the increasing frequency and intensity of forest fires in Bolivia, there is a lack of studies that directly analyze their impact on the health of the communities closest to these events. Most research has focused on environmental consequences, such as soil degradation and biodiversity loss, without assessing how prolonged exposure to smoke and other pollutants affects local populations. This research is essential because it allows us to understand the adverse effects on the health of people living in affected areas, providing scientific evidence to guide the implementation of prevention and mitigation strategies in public health.

## METHODOLOGY

### Population and study area

This cross-sectional study was carried out in San José de Chiquitos, department of Santa Cruz, Bolivia, in September 2024. San José de Chiquitos is located east of the capital of the department of Santa Cruz and has approximately 29,000 inhabitants, according to the 2012 census. The study was carried out with two communities that are located within the municipality of San José de Chiquitos, Los Ciros and Pororó, which belong to the Central de Comunidades Indígenas de Chiquitos Turubó. All the people who were treated in the brigade were included.

### Environmental Data Analysis

Data for the environmental analysis were obtained from ^1^. This data was used to create the maps (Figure 1) through ArcGIS online software. The graph (Figure 2) was made with the same data using the Prism v10 software.

**Figure 1.**
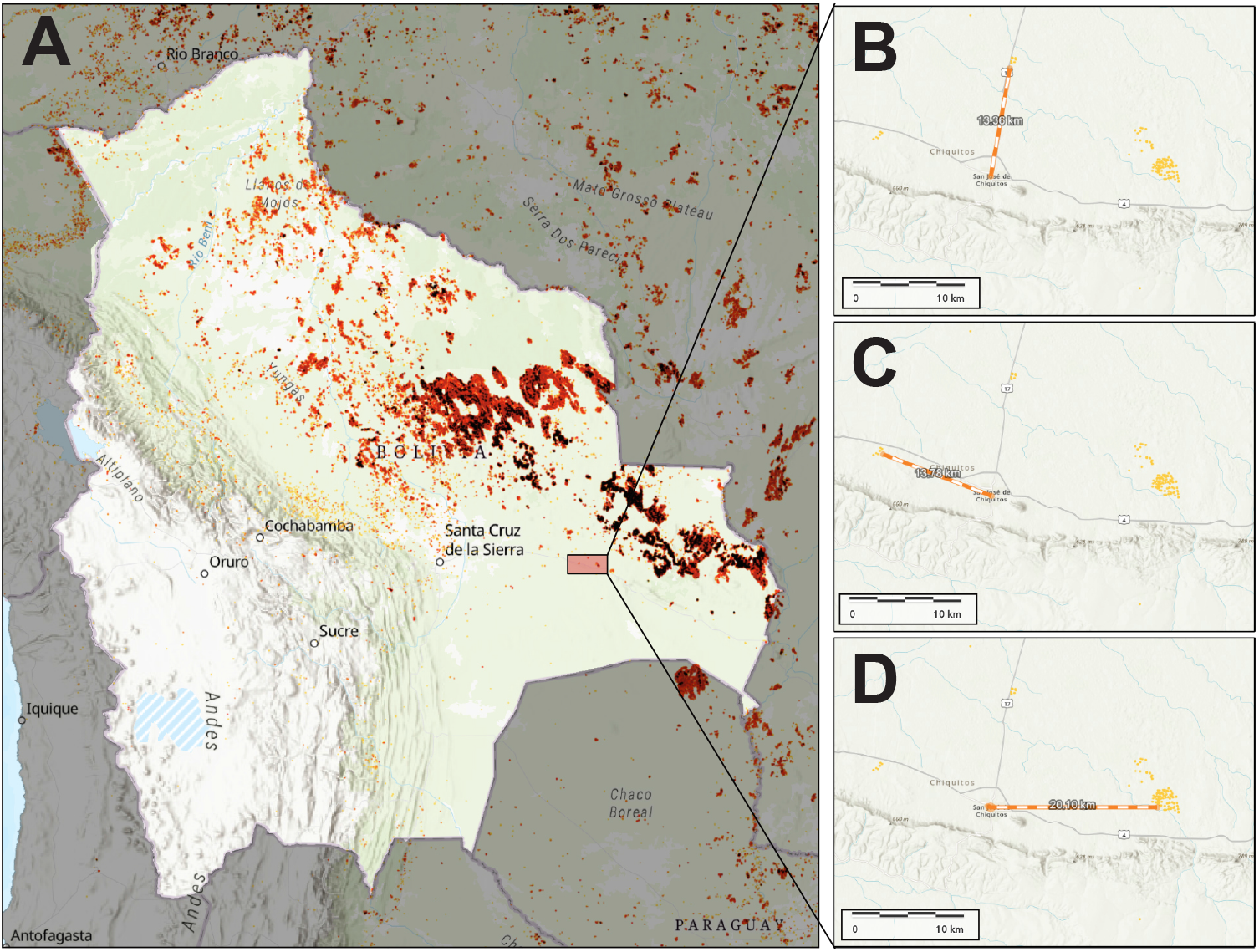
Forest fires in Bolivia during September 2024. **A**. Active heat sources throughout the national territory during September 2024. The rectangle shows the community of San José de Chiquitos. **B**. Active heat sources are 13.36 km north of the capital of San José de Chiquitos. **C**. Active heat sources are 13.78 km west of the capital of San José de Chiquitos. **D**. Active heat sources are 20.10 km east of the capital of San José de Chiquitos. Source of the data (Global Forest Watch, 2024).

**Figure 2.**
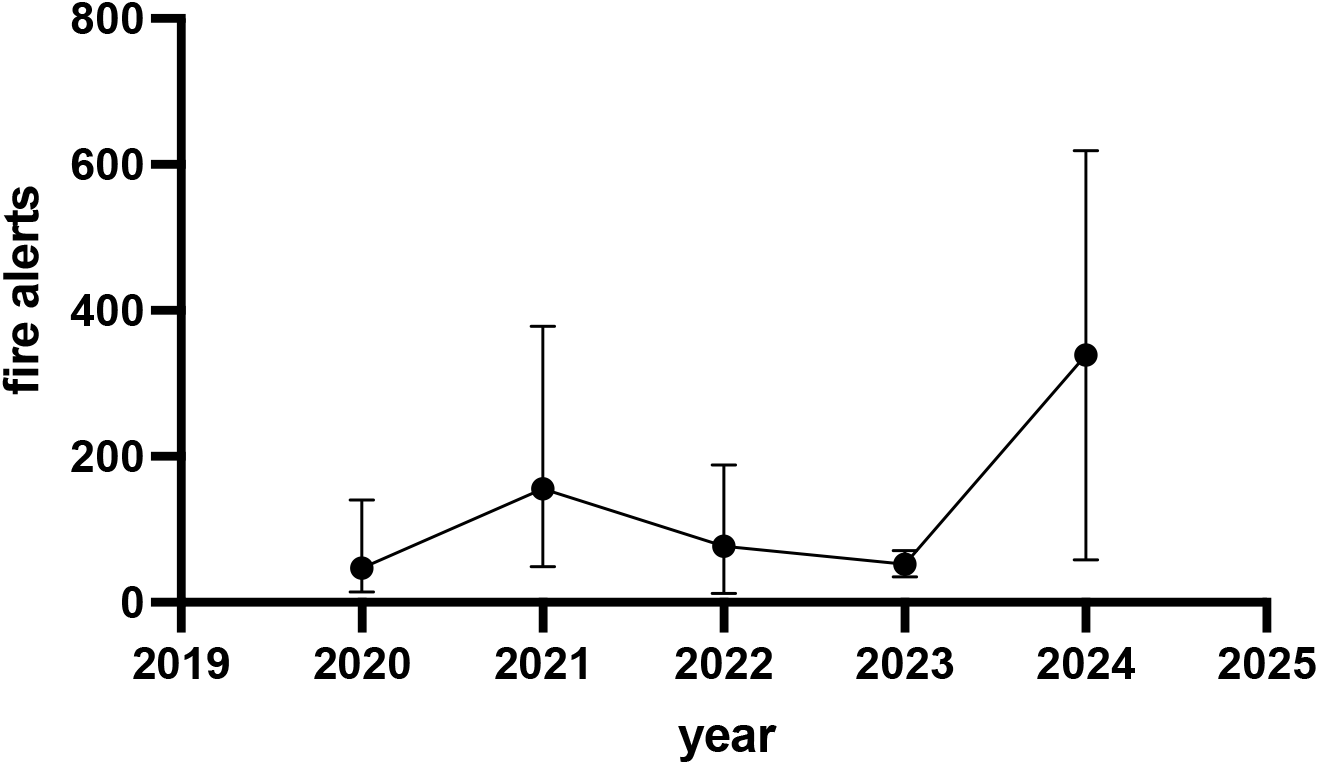
Trend of fire alerts in San José de Chiquitos in recent years. The figure shows the averages of fire trends in the last 4 years in San José de Chiquitos, with a significant increase in 2024. Data Source (Global Forest Watch, 2024)

### Demographic data collection and analysis

The participants’ data were obtained through the collection of epidemiological information obtained in a medical brigade carried out by the medical career of the Bolivian Catholic University “San Pablo” in the municipality of San José de Chiquitos. These data included information on pre-existing diseases, effects of smoke exposure, and methods of care established by the community members. These data were collected by professors and students in the university’s medical career.

The data analyses depended on the type of variables involved; for categorical variables, the Chi-square test was used, and for numerical variables, the median and Mann-Whitney test was used. All statistical analyses were performed in the Stata v.18 software.

### Ethical considerations

The present study obtained approval from the Bioethics Committee of the Bolivian Catholic University “San Pablo” to analyze secondary data. All participants verbally accepted the data collection in the epidemiological file, which did not contain sensitive or personal information.

## RESULTS

### Evolution of forest fires in Santa Cruz

In recent years, Bolivia has experienced a significant increase in the incidence and severity of forest fires, affecting various areas of forests and nature reserves protected by the state. Between 2010 and 2015, the Department of Santa Cruz accounted for 76% of forest loss due to forest fires, which increased to 86% from 2016 to 2022 ^10^. In 2019, Bolivia suffered large-scale fires that devastated approximately 6 million hectares of forests and savannahs, with the department of Santa Cruz being one of the most affected ^2^.

The situation worsened in 2024 when the country faced one of the worst fire seasons in its history. From May to October of that year, more than 36,800 heat sources were registered, 97% of which were concentrated in the departments of Beni and Santa Cruz. These fires devastated more than 10 million hectares, of which 58% corresponded to forests and 42% to other vegetation cover ^9^. The leading causes of these fires include agricultural practices such as clearing, which are used to prepare land for planting, which often gets out of control and causes large fires. In addition, policies that promote the expansion of the agricultural frontier have contributed to increased deforestation and vulnerability to fires. ^8^. This increase in the frequency and extent of forest fires in Bolivia, particularly in Santa Cruz, underscores the need to implement sustainable land management strategies and stricter environmental policies to mitigate future disasters. Figure 1 shows the geographical distribution of forest fires in the San José de Chiquitos region, showing an increase in the extension of the affected areas in recent years. Figure 2 shows the trend of fire alerts between 2019 and 2025, highlighting a significant increase in the number of alerts, especially in the years 2020 and 2023, with a peak recorded in 2023, indicating an intensification of forest burning activity in the region.

### Demographic and health characteristics of the study population in San José de Chiquitos

Table 1 summarizes the demographic characteristics of the population studied (N=35), with a predominance of women (65.7%) and a mean age of 39.77±19.67 years. Most participants are not engaged in agriculture (81.8%) and report not having underlying diseases (71.4%). Among pre-existing diseases, high blood pressure was the most common (14.3%). Table 2 shows significant differences in some health parameters between Los Ciros and Pororó communities. The average weight was higher in Los Ciros (73.27 kg) compared to Pororó (63.31 kg) (p=0.042). Likewise, significant differences were recorded in heart rate (p=0.003) and systolic pressure (p<0.001), being higher in Los Ciros.

**Table 1.** Demographic characteristics of the study population (N=35)

|  | Community |  |  |
| --- | --- | --- | --- |
|  | Los Ciro (16) | Pororó (19) | Total (35) |
| **Sex** |  |  |  |
| Man | 6 (37.5%) | 6 (31.6%) | 12 (34.3%) |
| Woman | 10 (62.5%) | 13 (68.4%) | 23 (65.7%) |
| **Age (years)** | 38.188 (18.534) | 41.105 (20.994) | 39.771 (19.674) |
| **He is a farmer** |  |  |  |
| No | 13 (86.7%) | 14 (77.8%) | 27 (81.8%) |
| Yes | 2 (13.3%) | 4 (22.2%) | 6 (18.2%) |
| **Time in the community (days)** | 22.929 (18.214) | 24.508 (14.067) | 23.771 (15.865) |
| **Underlying disease** |  |  |  |
| No | 11 (68.8%) | 14 (73.7%) | 25 (71.4%) |
| Asthma | 1 (6.2%) | 0 (0.0%) | 1 (2.9%) |
| Diabetes | 1 (6.2%) | 1 (5.3%) | 2 (5.7%) |
| Rheumatic fever | 1 (6.2%) | 0 (0.0%) | 1 (2.9%) |
| Hypertension | 2 (12.5%) | 3 (15.8%) | 5 (14.3%) |
| Hypertension, diabetes, and arrhythmia | 0 (0.0%) | 1 (5.3%) | 1 (2.9%) |

**Table 2.** Underlying diseases and health status.

|  | Community |  |  | P value |
| --- | --- | --- | --- | --- |
|  | Los Ciro (16) | Pororó (19) | Total (35) |  |
| Weight (kg) | 73.27 (16.298) | 63.31 (8.875) | 68.13 (13.745) | 0.042 |
| Height (cm) | 152.80 (31.260) | 161.50 (5.993) | 157.55 (21.586) | 0.255 |
| Temperatur (°C) | 35.82 (0.452) | 35.70 (.) | 35.81 (0.428) | 0.804 |
| Heart rate | 81.00 (6.587) | 72.64 (7.099) | 76.82 (7.954) | 0.003 |
| Systolic pressure (mmHg) | 134.79 (12.945) | 113.93 (12.431) | 124.36 (16.367) | <0.001 |
| Diastolic pressure (mmHg) | 79.21 (14.911) | 79.43 (11.022) | 79.32 (12.867) | 0.966 |

*All statistical analyses were performed using the median and Mann-Whitney test*

### Health Effect of Wildfire Smoke Exposure

Table 3 details the conditions associated with exposure to smoke. Respiratory distress was the most reported symptom (71.4%), followed by mucosal irritation (77.1%), sore throat (80.0%), and eye irritation (85.7%). Significant differences were observed in the presence of sore throat between communities (p=0.007), being more prevalent in Los Ciros. Table 4 describes the measures adopted by the population to mitigate the effects of smoke. The use of masks was the most frequent measure (63.6%), followed by staying indoors (54.5%) and the use of wet cloths (21.2%). Significant differences were found in the type of water consumed, highlighting a higher use of well water in Los Ciros (62.5%) compared to Pororó (21.1%) (p=0.013). Supplementary Tables 1 and 2 explore the relationship between care measures and the occurrence of respiratory diseases and irritations. No statistically significant associations were observed between using masks wet cloths, or staying indoors and reducing respiratory symptoms or irritation. However, drinking water consumption remained high in all categories, which could have partially helped to mitigate some adverse effects.

**Table 3.**
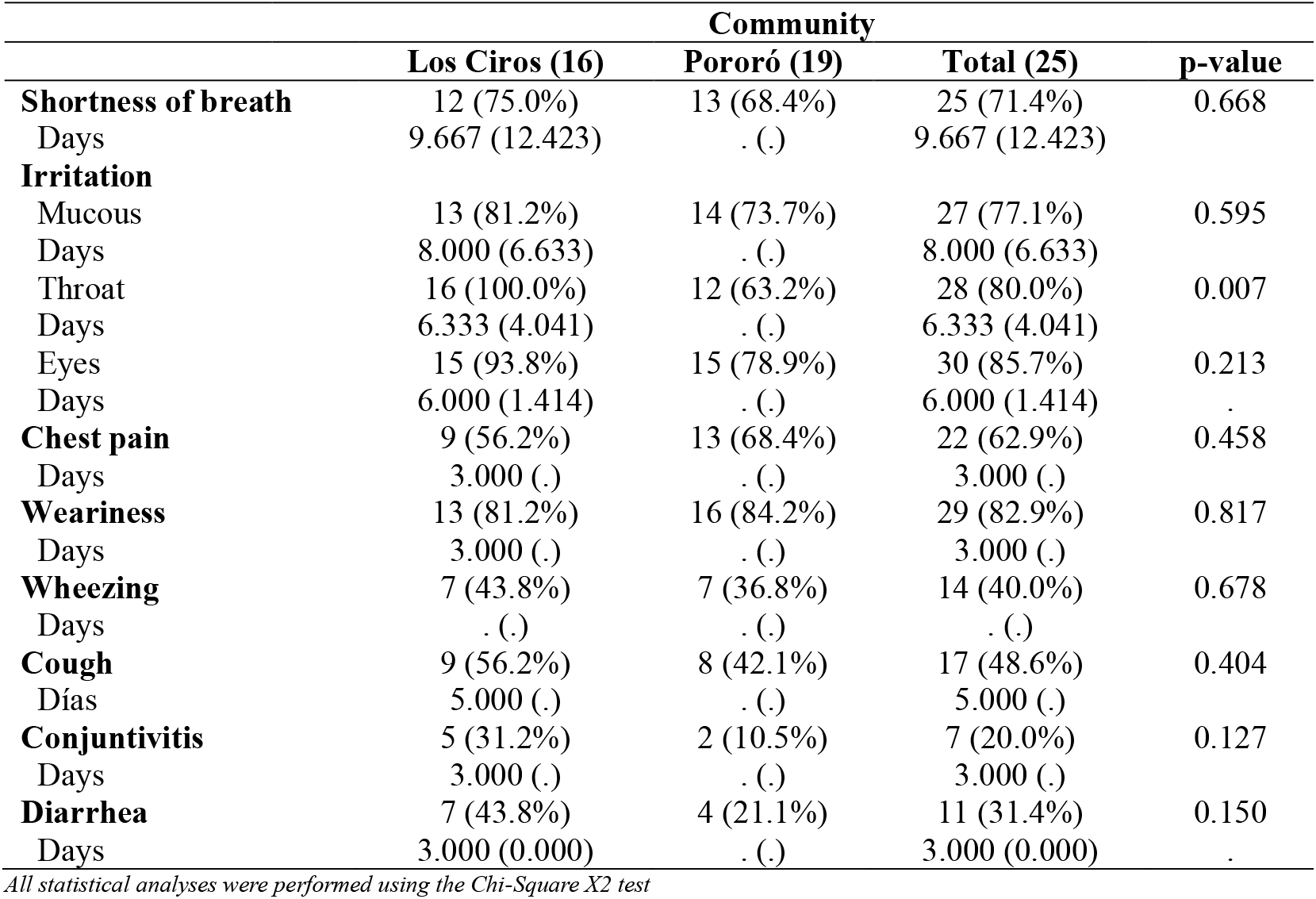
Conditions due to exposure to smoke.

*All statistical analyses were performed using the median and Mann-Whitney test*
| **Table 3. Conditions due to exposure to smoke** |  |  |  |  |
| --- | --- | --- | --- | --- |
|  | **Community** |  |  |  |
|  | **Los Ciro (16)** | **Pororó (19)** | **Total (25)** | **p-value** |
| **Shortness of breath** | 12 (75.0%) | 13 (68.4%) | 25 (71.4%) | 0.668 |
| Days | 9.667 (12.423) | . (.) | 9.667 (12.423) |  |
| **Irritation** |  |  |  |  |
| Mucous | 13 (81.2%) | 14 (73.7%) | 27 (77.1%) | 0.595 |
| Days | 8.000 (6.633) | . (.) | 8.000 (6.633) |  |
| Throat | 16 (100.0%) | 12 (63.2%) | 28 (80.0%) | 0.007 |
| Days | 6.333 (4.041) | . (.) | 6.333 (4.041) |  |
| Eyes | 15 (93.8%) | 15 (78.9%) | 30 (85.7%) | 0.213 |
| Days | 6.000 (1.414) | . (.) | 6.000 (1.414) | . |
| **Chest pain** | 9 (56.2%) | 13 (68.4%) | 22 (62.9%) | 0.458 |
| Days | 3.000 (.) | . (.) | 3.000 (.) |  |
| **Weariness** | 13 (81.2%) | 16 (84.2%) | 29 (82.9%) | 0.817 |
| Days | 3.000 (.) | . (.) | 3.000 (.) |  |
| **Wheezing** | 7 (43.8%) | 7 (36.8%) | 14 (40.0%) | 0.678 |
| Days | . (.) | . (.) | . (.) |  |
| **Cough** | 9 (56.2%) | 8 (42.1%) | 17 (48.6%) | 0.404 |
| Días | 5.000 (.) | . (.) | 5.000 (.) |  |
| **Conjunctivitis** | 5 (31.2%) | 2 (10.5%) | 7 (20.0%) | 0.127 |
| Days | 3.000 (.) | . (.) | 3.000 (.) |  |
| **Diarrhea** | 7 (43.8%) | 4 (21.1%) | 11 (31.4%) | 0.150 |
| Days | 3.000 (0.000) | . (.) | 3.000 (0.000) | . |

*All statistical analyses were performed using the Chi-Square X2 test*

**Table 4.** Care taken by community members to mitigate the effects of smoke.

*All statistical analyses were performed using the Chi-Square X2 test*
|  |  |  |  |  |
| --- | --- | --- | --- | --- |
|  | **Community** |  |  |  |
|  | **Los Ciro (16)** | **Pororó (19)** | **Total (35)** | **P value** |
| **Mask** | 11 (73.3%) | 10 (55.6%) | 21 (63.6%) | 0.290 |
| **Wet cloths** | 4 (28.6%) | 3 (15.8%) | 7 (21.2%) | 0.375 |
| **Staying indoors** | 8 (57.1%) | 10 (52.6%) | 18 (54.5%) | 0.797 |
| **Special Equipment** | 0 (0.0%) | 0 (0.0%) | 0 (0.0%) |  |
| **Water consumption** |  |  |  |  |
| Drinkable | 16 (100.0%) | 17 (89.5%) | 33 (94.3%) | 0.181 |
| Well | 10 (62.5%) | 4 (21.1%) | 14 (40.0%) | 0.013 |
| River | 0 (0.0%) | 0 (0.0%) | 0 (0.0%) |  |
| Tank | 7 (46.7%) | 0 (0.0%) | 7 (20.6%) | <0.001 |
| Bottle | 9 (56.2%) | 2 (10.5%) | 11 (31.4%) | 0.004 |

## DISCUSSION

Forest fires have increased in frequency and intensity in Bolivia in recent years, and 2024 has been particularly critical, with a devastated area greater than previous records ^2,6,7^. A study in 2023 established that soils are strongly affected by fires, reducing their fertility and production level ^5^. An article highlights that exposure to air pollutants from forest fires is associated with respiratory and cardiovascular problems and, specifically, damage to human cells’ genetic material ^11^. This phenomenon has generated a high risk to the health of the population exposed to smoke, especially in rural communities such as Losirios de San José de Chiquitos. The Pan American Health Organization reported that Santa Cruz was the department most affected by fires in 2024 ^6^, and San José de Chiquitos was one of the municipalities with the highest number of hectares burned in the same year ^7^. In this study, we evaluated the impact of air pollution from fires on the population’s health, finding that the adverse effects are mainly concentrated in respiratory and irritation symptoms, with variations depending on the baseline health status and the mitigation strategies adopted.

An essential finding of this study is that most participants were not farmers, nor did they have a high prevalence of underlying diseases, which is relevant in a rural population. However, Los Cirios presented a significantly different baseline state of health than Pororó. Los Ciros had a substantially higher weight and a higher prevalence of underlying diseases, although not significant, greater than that of Pororó. This shows that both populations, despite being neighbors, had different behaviors in their health. It is known that populations with a history of underlying diseases, mainly respiratory diseases, are usually more vulnerable to air pollution ^12,13^. These differences could be related to lifestyle habits, nutritional factors, or access to health services in each community, which could explain a greater susceptibility to the effects of smoke.

The effects of exposure to wildfire smoke were predominantly respiratory, followed by symptoms of irritation in mucous membranes, throat, and eyes in both communities. Although not all differences were statistically significant, a higher prevalence of symptoms was observed in Los Ciros compared to Pororó. This suggests that poorer baseline health may aggravate the effects of smoke exposure, while better health status may offer some protection ^4^. Previous studies have shown that air pollution from delicate particulate matter (PM2.5) and other compounds derived from biomass burning is directly linked to exacerbations of respiratory and cardiovascular diseases ^4,12–14^.

The mitigation strategies most used by the population were using masks, staying indoors, and using damp cloths. Although these measures can provide some protection, their effectiveness is limited in scenarios of prolonged exposure and high concentrations of pollutants ^3,15^. An interesting finding of this study was the significant difference in the type of water consumed, with higher use of well water in Los Ciros than in Pororó. Access to safe water sources is essential in situations of environmental crisis, as consuming contaminated water can aggravate the adverse effects on health ^3,16^. Supplementary Tables 1 and 2 analyzed the relationship between mitigation strategies and the occurrence of respiratory diseases and irritations without finding statistically significant associations between using masks, wet cloths, or staying indoors and reducing symptoms. This could be explained by the high concentration of pollutants and prolonged exposure to smoke, which exceeds the protective capacity of these strategies. However, drinking water consumption remained elevated in all categories, which could have helped mitigate some adverse effects.

The results of this study underscore the need to strengthen strategies for preventing and mitigating the health impact of forest fires. It is recommended that access to more effective protective equipment, such as special masks, be improved and air quality monitoring protocols in affected communities be established. In addition, it is essential to promote access to safe water sources and strengthen medical care in vulnerable areas. Given that forest fires in Bolivia have been on the rise, it is crucial to implement environmental policies that regulate the expansion of the agricultural frontier and reduce deforestation, which are the main factors contributing to these events’ recurrence. One of the limitations of this study was the lack of measurement of air quality in both communities, which could be decisive for the level of development of the effects produced by exposure to smoke. However, since these communities are neighbors, and due to the tremendous environmental pollution throughout the department, it is believed that both communities had the same level of exposure to smoke. Another limitation was the low sample size, which may have influenced our statistical analyses. However, this study intends to describe how smoke from forest fires affects the most vulnerable populations. In future research, it would be helpful to assess biomarkers of exposure in the affected population and to analyze the long-term effects of air pollution on health.

## Data Availability

All data produced in the present study are available upon reasonable request to the authors

## ACKNOWLEDGEMENTS

We thank the San José de Chiquitos Turubó community for helping us develop this research. We also thank the professors and students of medical careers who participated in the medical brigade and epidemiological data collection for this study. Finally, we would like to thank Drs. Adriana Amelunge and Mariana Santa Cruz for their collaboration and support during the development of this study.

## SUPPLEMENTARY TABLES

**Supplementary Table 1.** Relationship between care and respiratory diseases.

|  | Respiratory diseases |  |  | Total (38) | p-value |
| --- | --- | --- | --- | --- | --- |
|  | None (3) | 1-3 (20) | >3 (15) |  |  |
| **Mask** | 1 (33.3%) | 14 (70.0%) | 8 (61.5%) | 23 (63.9%) | 0.456 |
| **Wet cloths** | 0 (0.0%) | 5 (25.0%) | 3 (23.1%) | 8 (22.2%) | 0.621 |
| **Staying indoors** | 2 (66.7%) | 9 (45.0%) | 8 (61.5%) | 19 (52.8%) | 0.572 |
| **Special Equipment** | 0 (0.0%) | 0 (0.0%) | 0 (0.0%) | 0 (0.0%) |  |
| **Water consumption** |  |  |  |  |  |
| Drinkable | 3 (100.0%) | 20 (100.0%) | 13 (86.7%) | 36 (94.7%) | 0.198 |
| Well | 0 (0.0%) | 6 (30.0%) | 8 (53.3%) | 14 (36.8%) | 0.142 |
| River | 0 (0.0%) | 0 (0.0%) | 0 (0.0%) | 0 (0.0%) |  |
| Tank | 0 (0.0%) | 4 (21.1%) | 4 (26.7%) | 8 (21.6%) | 0.590 |
| Bottle | 0 (0.0%) | 6 (30.0%) | 5 (33.3%) | 11 (28.9%) | 0.503 |

All p-values were obtained using Pearson's chi-squared assay

**Supplementary Table 2.**
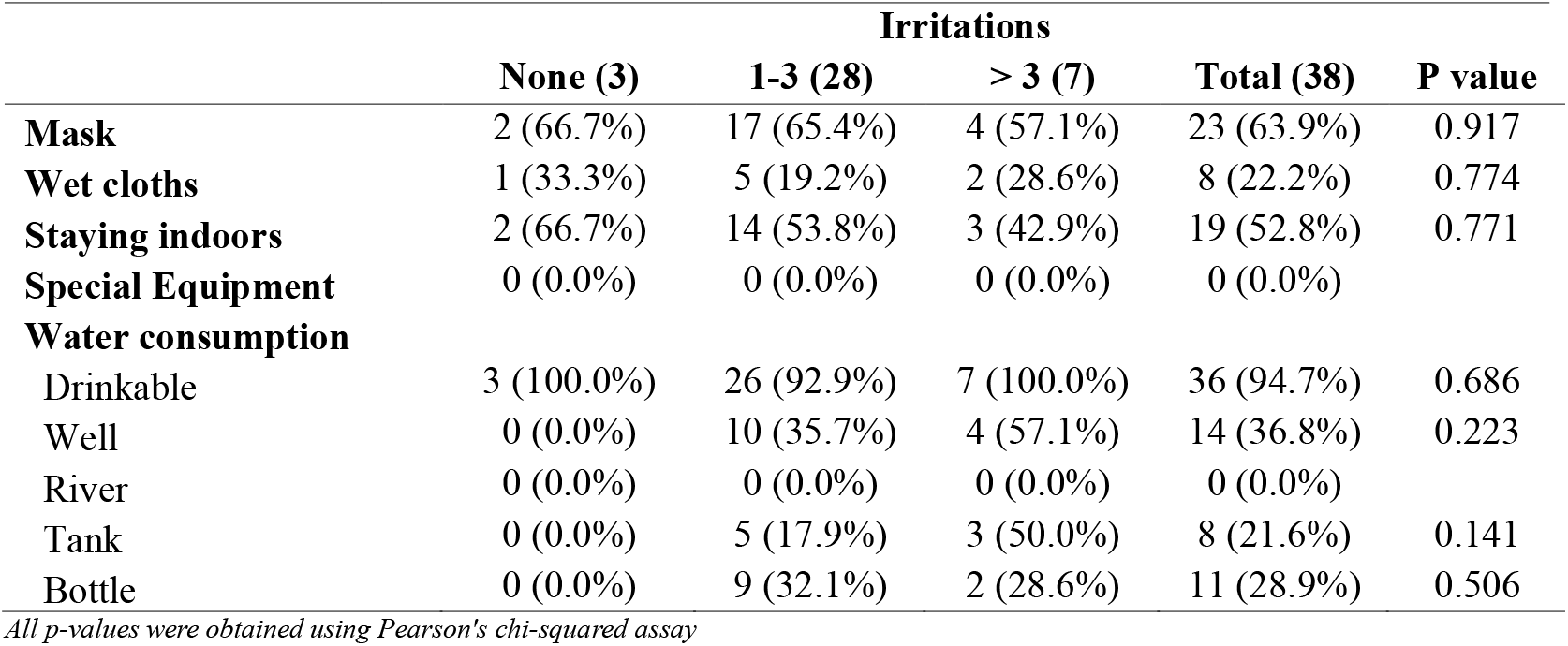
Relationship between care and irritation.

|  | Irritations |  |  | Total (38) | P value |
| --- | --- | --- | --- | --- | --- |
|  | None (3) | 1-3 (28) | > 3 (7) |  |  |
| **Mask** | 2 (66.7%) | 17 (65.4%) | 4 (57.1%) | 23 (63.9%) | 0.917 |
| **Wet cloths** | 1 (33.3%) | 5 (19.2%) | 2 (28.6%) | 8 (22.2%) | 0.774 |
| **Staying indoors** | 2 (66.7%) | 14 (53.8%) | 3 (42.9%) | 19 (52.8%) | 0.771 |
| **Special Equipment** | 0 (0.0%) | 0 (0.0%) | 0 (0.0%) | 0 (0.0%) |  |
| **Water consumption** |  |  |  |  |  |
| Drinkable | 3 (100.0%) | 26 (92.9%) | 7 (100.0%) | 36 (94.7%) | 0.686 |
| Well | 0 (0.0%) | 10 (35.7%) | 4 (57.1%) | 14 (36.8%) | 0.223 |
| River | 0 (0.0%) | 0 (0.0%) | 0 (0.0%) | 0 (0.0%) |  |
| Tank | 0 (0.0%) | 5 (17.9%) | 3 (50.0%) | 8 (21.6%) | 0.141 |
| Bottle | 0 (0.0%) | 9 (32.1%) | 2 (28.6%) | 11 (28.9%) | 0.506 |

All p-values were obtained using Pearson's chi-squared assay

